# Feasibility of Vibrotactile P50 Sensory-Gating Assessment Using Portable EEG

**DOI:** 10.64898/2026.11.06.26367020

**Authors:** Jean-Paul Badjo, Fow-Sen Choa

## Abstract

Sensory gating (SG), most commonly quantified using the auditory P50 event-related potential elicited by a paired “double-click” paradigm, is a well-established biomarker of early sensory inhibition that reflects the brain’s ability to suppress responses to repetitive sensory input. Although auditory SG paradigms have been widely used in both research and clinical settings, they impose an unavoidable auditory load and are generally limited to controlled laboratory environments, reducing their suitability for ecological monitoring and applications in which auditory resources must remain available. This study evaluated the feasibility of a portable vibrotactile sensory-gating system designed to assess inhibitory processing without engaging the auditory or visual modalities. Fifteen healthy adult participants completed both auditory and vibrotactile paired-stimulus conditions using a within-subject experimental design. Vibrotactile stimuli were delivered through transducers positioned on the volar surface of the forearm while electroencephalographic (EEG) activity was recorded using a 32-channel Emotiv EPOC Flex system. Event-related responses were extracted and analyzed using conventional sensory-gating metrics to enable direct comparison between modalities. Vibrotactile stimulation elicited time-locked P50 responses with attenuation of S2 relative to S1. Mean S2/S1 ratios were 0.521 ± 0.041 for the auditory condition and 0.524 ± 0.064 for the vibrotactile condition. The paired difference was 0.0029 (95% CI, −0.0221 to 0.0279), t(14) = 0.249, p = 0.807, dz = 0.064. No statistically significant difference was detected between modalities in this sample. These group-level findings support further evaluation of portable vibrotactile P50 measurement.

## I. Introduction

Sensory gating (SG) is a neurophysiological mechanism that attenuates responses to repeated or irrelevant sensory input [1]. Impaired auditory P50 gating is especially well documented in schizophrenia, although its clinical and cognitive correlates remain incompletely established [2-5,10]. Altered P50 gating has also been reported in adults with ADHD, bipolar disorder, and major depressive disorder [6-8]. Findings in autism spectrum disorder are component- and sample-dependent; one study found N100 rather than P50 group differences [9].

A common method for evaluating SG is the paired-stimulus, or “double-click,” paradigm, in which two identical stimuli (S1 and S2) are presented in rapid succession [1,3]. Under healthy conditions, the neural response to S2 is attenuated relative to S1. Sensory gating is commonly quantified as S2/S1, with higher ratios indicating weaker suppression [1,3]. Auditory P50 gating has been studied extensively in schizophrenia [2-5]. N100 and P200 gating have also been examined and may relate to behavioral inhibition, attention, and working memory [2,10]. Separately, paired-pulse TMS studies have reported reduced short-interval intracortical inhibition in schizophrenia; this is a distinct inhibitory measure rather than a P50 measure [11].

Although auditory P50 paradigms remain the most widely accepted approach for evaluating SG, they possess several practical limitations that restrict their use outside controlled laboratory environments. Auditory stimulation necessarily occupies the auditory channel and may interfere with communication, environmental awareness, and concurrent tasks. Furthermore, conventional SG assessments are typically conducted during brief experimental sessions, limiting their ability to capture fluctuations in inhibitory function that occur over time. Given that attention, stress, fatigue, and cognitive workload vary throughout daily life, there is increasing interest in developing portable neurophysiological tools capable of longitudinal monitoring in naturalistic settings.

Vibrotactile stimulation can elicit measurable somatosensory cortical responses, while attention, movement, stimulus repetition, and sensory context can modulate somatosensory evoked activity [12-20]. Because tactile stimulation does not intentionally occupy the visual channel and can leave the auditory channel available, it warrants evaluation as an alternative paired-stimulus modality.

Portable and lower-cost EEG systems have supported mobile recordings and auditory ERP measurement [21-23]. Validation studies of Emotiv EPOC systems support their use for several EEG and ERP measures; EPOC Flex has also been compared with a research-grade system, and dedicated timing studies show that event-marker lag must be measured and corrected [22-26]. Portable EEG and tactile stimulation may therefore support future repeated assessment outside conventional laboratories.

Accordingly, the immediate question was whether vibrotactile stimulation could produce measurable P50 suppression in healthy adults under controlled laboratory conditions.

This study evaluated the laboratory feasibility of recording vibrotactile P50 sensory-gating responses with the Emotiv EPOC Flex and compared participant-level S2/S1 ratios with an auditory reference condition.

## II. Materials and Methods

### A. Experimental Design

Fifteen healthy adults aged 18-35 years participated (mean ± SD, 24.7 ± 4.1 years; eight men and seven women); all were right-handed. Exclusion criteria were self-reported neurological or psychiatric illness, traumatic brain injury with loss of consciousness, current psychoactive-medication use, uncorrected hearing impairment, or reduced tactile sensitivity. Participants were instructed to obtain at least 6 h of sleep and avoid caffeine and nicotine for at least 2 h before testing. Eligibility, medication use, recent caffeine and nicotine use, sleep duration, and current fatigue were confirmed before recording; hearing and tactile perception were confirmed during familiarization.

The auditory double-click paradigm was included as the established reference condition because auditory P50 suppression has been extensively characterized in the sensory-gating literature and is widely regarded as a reliable measure of inhibitory processing. In this paradigm, healthy individuals typically exhibit reduced neural responses to the second of two closely spaced stimuli, reflecting the brain’s ability to filter redundant sensory information. Replicating this phenomenon within the present cohort provided an internal benchmark against which the performance of the vibrotactile paradigm could be evaluated.

The vibrotactile paired-stimulus condition was developed to determine whether similar inhibitory mechanisms could be assessed through somatosensory stimulation without introducing additional auditory or visual demands. This distinction is particularly relevant for populations in which auditory processing may already be compromised or when experimental paradigms require the preservation of auditory and visual resources for concurrent tasks. By employing tactile stimulation, the present study sought to investigate whether sensory gating could be measured using an alternative modality while maintaining the characteristic suppression patterns observed in traditional auditory paradigms.

The within-subject design permits comparison of group means and exploratory cross-modal agreement.

### B. Vibrotactile Stimulation and EEG Acquisition

Vibrotactile stimulation was delivered with a forearm-mounted linear moving-magnet tactor secured to the volar forearm by an adjustable strap. The tactor was driven through an external amplifier, and each stimulus was a 10 ms, 250 Hz burst. Participants remained seated with the stimulated arm supported and relaxed, and the actuator and cables were secured away from EEG leads to minimize movement and electrical interference.

Stimulus timing and actuator control used an Arduino UNO R4 Minima with a 48 MHz Renesas RA4M1 Arm Cortex-M4 microcontroller and 5 V logic. The Arduino generated the 10 ms S1 and S2 drive pulses, maintained the 500 ms interstimulus interval and 8 s recovery interval, and issued the event-timing signal used for EEG alignment.

Participants wore the same closed-back HyperX Cloud III over-ear headset throughout both conditions. Auditory stimuli were delivered through the headset in the auditory condition; audio playback was disabled during vibrotactile trials.

EEG was recorded at 128 samples/s with a 32-channel Emotiv EPOC Flex using international 10-20 electrode positions. The manufacturer-specified acquisition bandwidth was 0.2-45 Hz with a built-in fifth-order sinc filter and high attenuation at 50 and 60 Hz. No additional notch filter was applied. The Emotiv CMS/DRL common-mode reference was used without offline re-referencing. Contact quality was monitored with EmotivPRO and required to remain in the manufacturer’s acceptable green range.

The recorded EPOC Flex montage included Fp1, Fp2, C3, Cz, and C4 within the manufacturer-documented 32-channel configuration. Cz was used for the prespecified P50 analysis. Figure S5 identifies the primary Cz channel and the auxiliary Fp1, Fp2, C3, and C4 locations within the montage.

All participants completed the auditory condition first and the vibrotactile condition second. Each trial included an 8 s post-S2 recovery interval followed by the 2 s prestimulus period of the next trial, producing approximately 10 s between S2 and the next S1. This interval exceeded the 4 s inter-trial interval previously shown to support recovery of somatosensory gating [27].

### C. Experimental Protocol

Each 10.52 s trial comprised a 2.000 s prestimulus period, a 10 ms S1 stimulus, a 500 ms interstimulus interval, a 10 ms S2 stimulus, and an 8 s recovery interval.

Each trial began with a 2.000 s prestimulus baseline period during which no experimental stimuli were delivered. This interval allowed ongoing EEG activity to stabilize and provided a reference period against which stimulus-evoked responses could be evaluated. Following the baseline period, the first stimulus (S1), consisting of either a 10 ms auditory pulse or a 10 ms vibrotactile pulse depending on the experimental condition, was presented from 2.000-2.010 s. S1 served as the conditioning stimulus intended to engage the neural mechanisms involved in sensory registration and inhibitory control.

After a 500 ms interstimulus interval (ISI), the second stimulus (S2) was delivered from 2.510-2.520 s. The 500 ms ISI was selected to maintain consistency with established sensory-gating protocols and to facilitate comparison with prior studies utilizing P50 suppression paradigms. Under typical gating conditions, the neural response elicited by S2 is reduced relative to the response generated by S1, reflecting the inhibitory processes responsible for filtering repetitive sensory input. The relative amplitudes of the responses to S1 and S2 therefore provide an index of sensory-gating performance.

The 8 s recovery interval reduced immediate carryover between trials. Participants remained seated and were instructed to minimize blinking, facial movement, and body movement.

Thirty-one trials were acquired per participant and modality (465 paired-stimulus trials per modality). The clear averaged waveforms in Figure 4 and the measured signal-to-noise ratios support the sufficiency of 31 trials for this feasibility analysis. Signal-to-noise estimates and within-participant bootstrap confidence intervals are reported in the Supplementary Materials.

**Figure 1.**
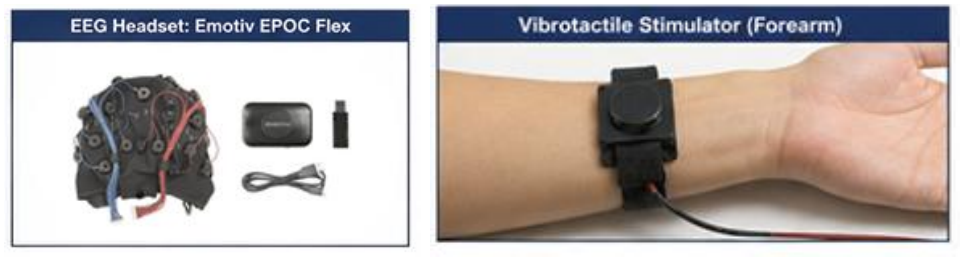
Portable vibrotactile sensory-gating measurement system and programmable controller platform. The system combines an Emotiv EPOC Flex 32-channel EEG headset, an Arduino UNO R4 Minima controller, an actuator-driver interface, and a forearm-mounted vibrotactile stimulator. The Arduino uses a 48 MHz Renesas RA4M1 Arm Cortex-M4 microcontroller with 5 V logic. Custom firmware generated the 10 ms S1 and S2 drive pulses, maintained the 500 ms interstimulus interval and 8 s recovery interval, and issued the event-timing signal used to align stimulus onset with the EEG recording. The integrated platform enables controlled paired vibrotactile stimulation and simultaneous EEG acquisition for assessment of P50 sensory gating.

**Figure 2.**
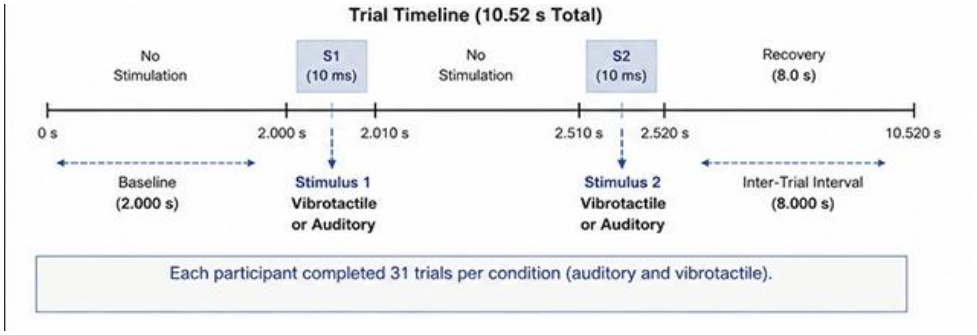
Experimental timing sequence for auditory and vibrotactile sensory-gating assessment. Each trial consisted of a 2.000 s prestimulus baseline period followed by two identical 10 ms stimuli (S1 and S2) separated by a 500 ms interstimulus interval. An 8 s recovery period followed S2 before initiation of the subsequent trial. EEG activity was recorded continuously throughout the protocol, and sensory-gating performance was quantified by comparing the P50 responses elicited by S1 and S2 using the conventional S2/S1 amplitude ratio.

**Figure 3.**
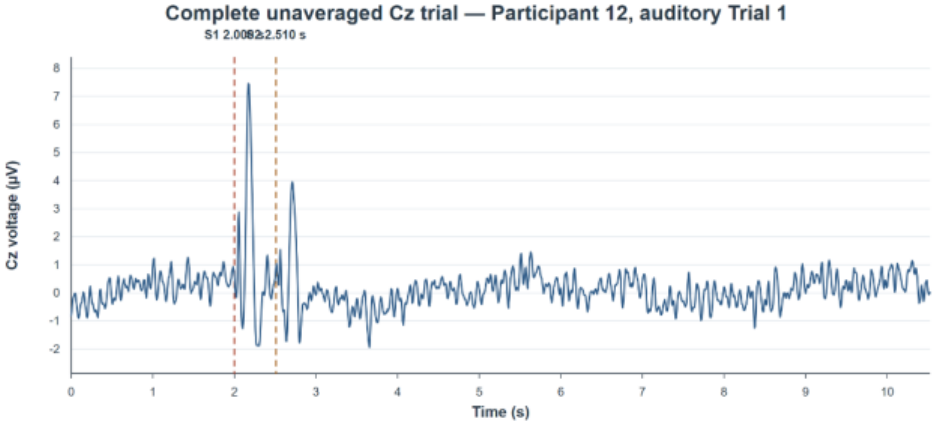
Original single-trial auditory Cz ERP from Participant 12, shown across the complete 10.52 s trial. The trace includes the 2.000 s prestimulus interval, the S1 onset at 2.000 s, the S2 onset at 2.510 s, and the subsequent 8 s recovery period. This unaveraged trace shows the scale and temporal context of the recorded EEG before participant-level averaging and P50-focused processing.

**Figure 4.**
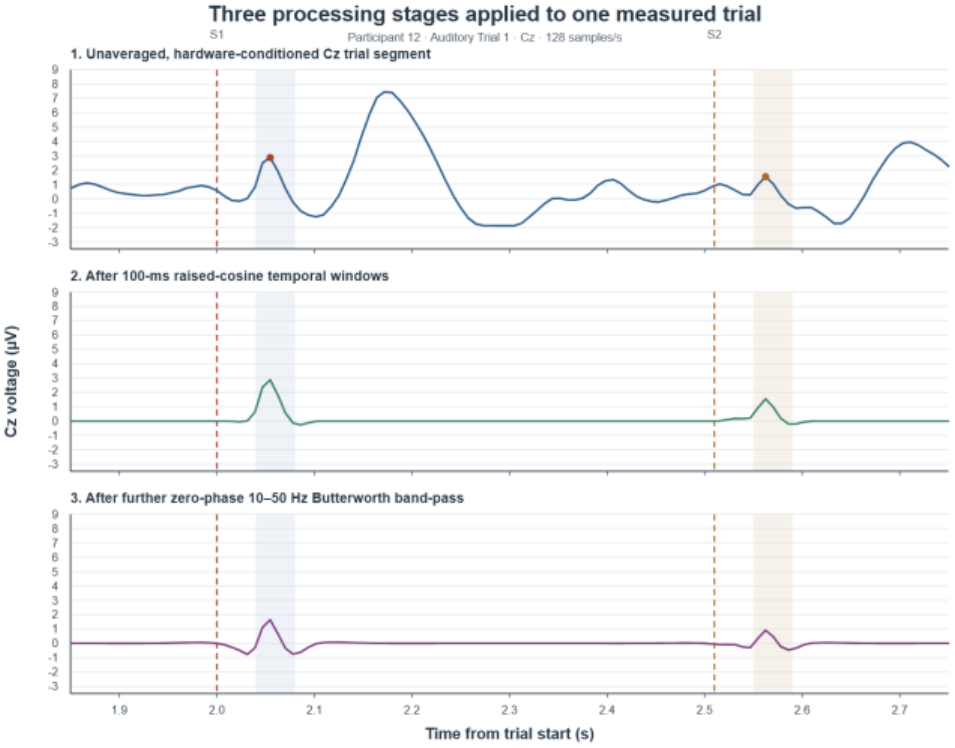
Participant 12 auditory Cz ERP signal-processing sequence. The ERP was obtained by separately averaging 31 baseline-corrected S1- and S2-locked trials using the −100 to 0 ms prestimulus interval. The participant-average waveform was then processed with a 100 ms single-cycle raised-cosine window centered on the detected P50 peak, followed by fourth-order zero-phase 10-50 Hz Butterworth band-pass filtering. Averaging reduced trial-specific background activity, windowing emphasized the P50 interval, and forward-backward filtering reduced slow drift and high-frequency noise without systematic phase delay.

### D. Stimulus Validation

Auditory stimuli were 10 ms, 1 kHz tone bursts with 1 ms cosine-squared onset and offset ramps, presented binaurally at 80 dB SPL through a wired HyperX Cloud III headset (model 727A8AA). The sequence was created in Adobe Premiere Pro, exported as one audio file, and played through Audacity.

Auditory output was calibrated at the earpiece with a sound-level meter and ear coupler. Across 100 pulses measured with a microphone, trigger-to-acoustic-onset latency was 18.6 ± 0.9 ms (peak-to-peak range, 3.2 ms) and pulse duration was 10.2 ± 0.4 ms. Event markers were shifted by 18.6 ms before epoching. OBS Studio recorded the playback, microphone signal, and EEG display as a synchronized validation record.

Vibrotactile timing was validated across 100 pulses using a two-channel digital oscilloscope and a piezoelectric contact sensor positioned at the actuator-skin interface. The first oscilloscope channel recorded the microcontroller drive signal, while the second channel simultaneously recorded the piezoelectric response. The resulting measurement therefore included the electrical-drive, actuator-motion, sensor-response, and electrical-conversion components of the validation chain.

Electrical onset was defined at the 50% point of the drive-signal rising edge. Mechanical onset was defined as the first sustained piezoelectric response exceeding five times the prestimulus baseline RMS. Rise and fall times were measured between the 10% and 90% response levels, and mechanical duration was measured between the rising and falling 10% crossings.

Trigger-to-mechanical-onset latency was 6.3 ± 0.8 ms, with a peak-to-peak onset range of 2.9 ms. Rise time was 2.1 ± 0.4 ms, fall time was 2.4 ± 0.5 ms, and mechanical pulse duration was 10.4 ± 0.7 ms. The corresponding EEG event marker occurred 1.2 ± 0.3 ms after the electrical trigger. Mechanical onset therefore occurred approximately 5.1 ms after the EEG marker, and this net delay was incorporated into vibrotactile epoch alignment.

During familiarization, participants rated perceived intensity on a 7-point scale. The auditory level remained at 80 dB SPL, and vibrotactile intensity was adjusted until both stimuli were detectable, comfortable, and within one scale point; settings then remained fixed.

An active off-skin sham condition also served as an electrical-coupling control. The same powered transducer, EEG system, cable configuration, stimulus timing, event markers, and processing procedure were retained, while the transducer was operated adjacent to the forearm without contacting the skin. The P50 observed during skin-contact stimulation was not reproduced in the off-skin sham condition. Because the electrical and cable configurations remained unchanged while only skin contact was removed, this result indicates that direct electrical coupling from the transducer drive system did not generate the recorded P50.

### E. EEG Processing and Statistical Analysis

EEG preprocessing was performed in EEGLAB [28]. Independent Component Analysis was applied to the continuous multichannel recordings before epoching to separate ocular and muscular artifact components from the EEG signal. Artifact-related components were removed, and the retained components were back-projected to the continuous channel data before segmentation.

After ICA correction, data were segmented separately around latency-corrected S1 and S2 onsets and baseline corrected by subtracting the mean voltage from −100 to 0 ms before each corresponding stimulus. Trial-level P50 amplitudes and S2/S1 ratios were calculated from the baseline-corrected sampled Cz epochs. To construct the participant-level waveforms, the 31 baseline-corrected Cz epochs were averaged separately for S1 and S2 within each participant. A 100 ms single-cycle raised-cosine temporal window centered on the detected P50 peak was applied to each participant average, and the windowed average was then filtered from 10 to 50 Hz with a fourth-order Butterworth filter using zero-phase forward-backward filtering (scipy.signal.filtfilt).

The 10 Hz high-pass cutoff attenuated slow baseline fluctuations and other low-frequency activity outside the P50-focused analysis band, while the 50 Hz low-pass cutoff attenuated higher-frequency noise. The Butterworth design provided a smooth passband without ripple. Applying the filter once in the forward direction and again in the reverse direction canceled phase delay, thereby avoiding systematic temporal displacement of the P50 response.

The 128 samples/s rate corresponds to 7.8125 ms per sample and a 64 Hz Nyquist frequency. The 40-80 ms search interval therefore contained approximately five to six sample locations. Analyses emphasized amplitude and S2/S1 ratio rather than precise latency, consistent with general ERP and neural time-series guidance concerning sampling density, filtering, and peak estimation [29,30].

Electrode contact and signal quality were monitored continuously. Recording began only after acceptable indicators were obtained. If contact loss, unacceptable quality, or acquisition interruption occurred, the headset was adjusted and the recording was restarted; incomplete recordings were not combined or counted.

### III. Equations

Here, s ∈ {S1, S2} identifies the stimulus; m ∈ {A, V} identifies the auditory or vibrotactile modality; i identifies the participant; j identifies the trial; r is the trial-level S2/S1 ratio; R is the participant-level mean of the 31 trial ratios; d_i_ is the paired participant difference calculated as vibrotactile minus auditory; SNR is the linear signal-to-noise ratio; and RMS is the root-mean-square prestimulus baseline voltage.

P50 was quantified at Cz using a predefined baseline-to-peak procedure. The 8 s recovery interval between trials reduced residual influence from the preceding paired-stimulus gating response before the prestimulus baseline was calculated. For every trial, S1 and S2 were measured independently as the maximum positive sampled voltage from 40 to 80 ms after the corresponding latency-corrected stimulus onset, following subtraction of the −100 to 0 ms baseline. The procedure used sampled values without manual peak selection, fractional-sample interpolation, minimum-prominence adjustment, peak-to-trough measurement, or temporal peak alignment. Trial-to-trial latency variability was handled by applying the same bounded search interval independently to each response. The same channel, baseline interval, 40-80 ms search interval, and peak-selection rule were applied consistently across all participants, trials, stimuli, and modalities.

Cz was selected a priori because P50 is commonly assessed over central/frontocentral scalp locations. The P50 analyses were restricted to Cz. The supplementary material provides the Cz waveforms and participant-level Cz S1, S2, and S2/S1 measurements used in the reported analyses.

A trial-level sensory-gating ratio was calculated as S2/S1, and each participant-level ratio was the arithmetic mean of the 31 trial-level ratios. Lower ratios indicate greater attenuation of the second response. The 31-trial averaged waveforms characterized ERP morphology, while participant-level ratios were calculated from the trial-level measurements.

To construct the participant-level waveforms, the 31 baseline-corrected Cz epochs were first averaged separately for S1 and S2 within each participant. A 100 ms single-cycle raised-cosine temporal window centered on the detected P50 peak was applied to each participant average. The windowed average was then filtered from 10 to 50 Hz with a fourth-order Butterworth filter using zero-phase forward-backward filtering (scipy.signal.filtfilt). For the group waveform, the resulting participant-level traces were temporally shifted so that their detected P50 peaks occurred at a common reference latency within 40-80 ms, with S1 and S2 aligned separately, and the 15 participant-level traces were averaged equally. This sequence produced the participant and group waveforms presented in Figures 4, 5, and S1.

**Figure 5.**
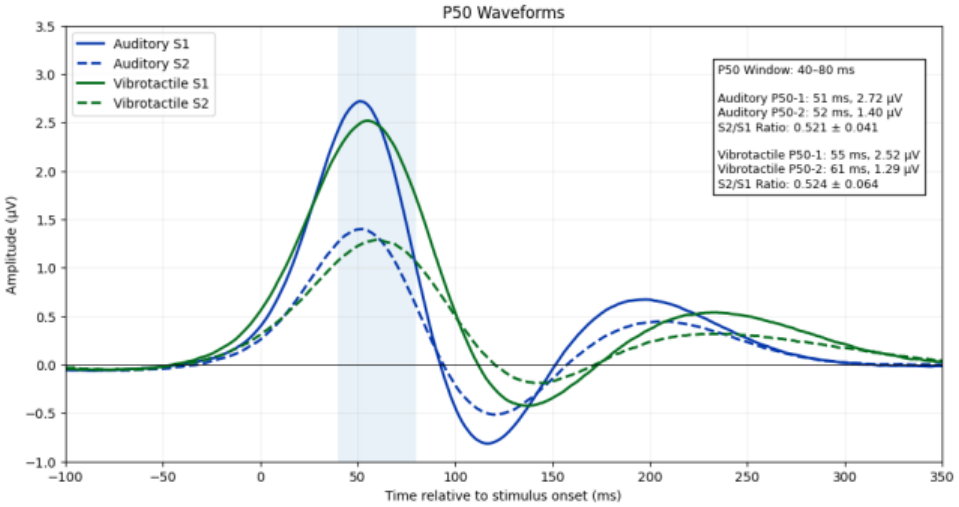
Processed auditory and vibrotactile Cz waveforms for the first (S1) and second (S2) stimuli. Each participant-level waveform was generated from the average of 31 baseline-corrected epochs, followed by a 100 ms single-cycle raised-cosine temporal window and a fourth-order zero-phase 10-50 Hz Butterworth band-pass filter. For the group waveform, participant-level P50 peaks were aligned to a common reference latency within 40-80 ms, with S1 and S2 aligned separately, and the 15 traces were averaged equally. The shaded 40-80 ms region denotes the predefined P50 measurement window. The annotated amplitudes, participant-level S2/S1 ratios, and statistical analyses report the predefined baseline-to-peak measurements described in Section 2.5.

**Figure 6.**
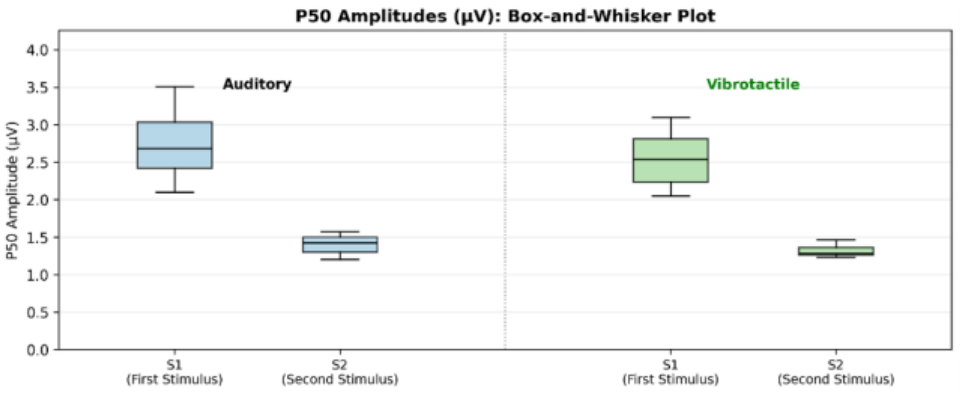
Subject-level box-and-whisker plots illustrating the distribution of auditory and vibrotactile P50-1 (S1) and P50-2 (S2) amplitudes across all 15 participants. The plots summarize the variability in stimulus-evoked responses observed within each modality, with boxes representing the interquartile range, central lines indicating the median amplitude, and whiskers depicting the overall spread of the data. Comparison of P50-1 and P50-2 amplitudes demonstrates the characteristic attenuation of the second response associated with sensory-gating mechanisms.

**Figure 7.**
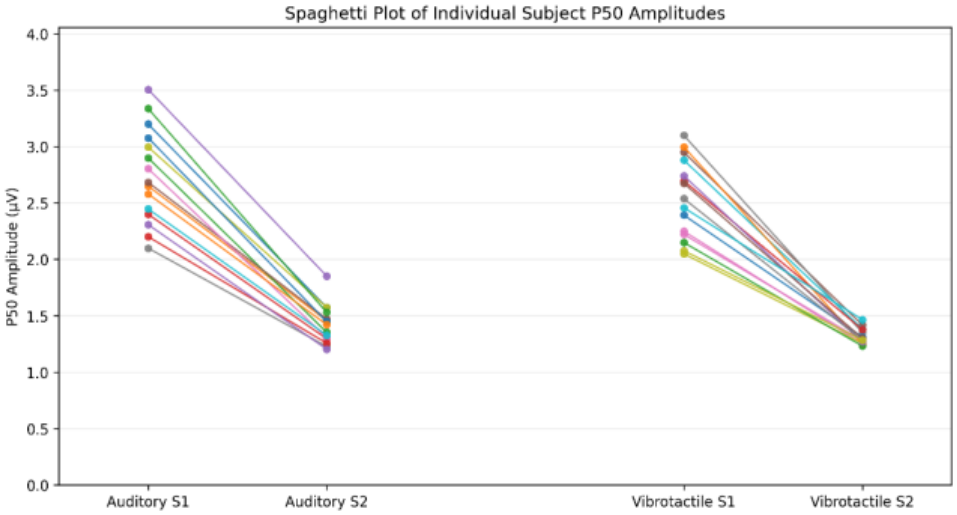
Subject-level spaghetti plots showing participant-specific changes in mean P50 amplitudes between the first stimulus (P50-1/S1) and second stimulus (P50-2/S2) for both auditory and vibrotactile conditions. Each connected pair of points corresponds to an individual participant, facilitating direct visualization of within-subject sensory-gating effects and inter-individual variability across modalities. Downward trajectories reflect attenuation of the second response, a characteristic feature of intact sensory gating.

Auditory and vibrotactile S2/S1 ratios were compared with a two-tailed paired-samples t-test. The paired difference was defined as vibrotactile minus auditory and reported with its 95% confidence interval, t statistic, two-sided p value, and paired effect size dz. Statistical significance was α = 0.05.

Statistical symbols are defined as follows: SD, standard deviation; CI, confidence interval; t(df), the t statistic with df degrees of freedom; p, the two-sided probability value; dz, the paired-samples standardized mean difference; α, the statistical significance threshold; r, Pearson correlation; ρ, Spearman rank correlation; and ICC(A,1), the two-way mixed-effects, absolute-agreement, single-measure intraclass correlation coefficient.

Exploratory individual-level agreement was assessed with Pearson and Spearman correlations, a two-way mixed-effects absolute-agreement single-measure intraclass correlation [ICC(A,1)], and Bland-Altman analysis. Potential proportional bias was tested by regressing paired differences on participant means.

Signal-to-noise ratio was the absolute P50 peak amplitude divided by prestimulus baseline RMS. Percentile 95% confidence intervals for participant ratios used 5,000 within-participant resamples of the 31 trial-level ratios with replacement. Analyses and visualizations used Python, NumPy, Pandas, SciPy, and Matplotlib.

Trial-number sensitivity analyses tested whether progressive habituation, fatigue, learning, or time-on-task affected the measurements across the 31 trials. Within each modality, participant-adjusted linear models regressed S1 amplitude, S2 amplitude, and the S2/S1 ratio on trial number. Trial-by-modality interaction models tested whether any trend differed between conditions. A paired early-versus-late comparison contrasted trials 1-10 with trials 22-31. Statistical significance was set at p < 0.05.

## IV. Results

### A. P50 Waveforms, Box-and-Whisker Plots, and Spaghetti Plots

Figure 3 presents the original unfiltered, non-peak-aligned single-trial Cz waveforms across the full 10.52 s trials, and Figure 4 documents each processing stage. Together, these figures show the full recorded temporal context and the resulting P50-focused morphology.

Figure 5 shows the processed group-mean P50 waveforms following the waveform-construction sequence described in Section 2.5. Participant-level P50 measurements were obtained using the predefined baseline-to-peak procedure described in Section 2.5.

Figure 8 presents the Bland-Altman comparison of participant-level auditory and vibrotactile S2/S1 ratios. The mean bias was 0.0029, and all participant differences fell within the 95% limits of agreement (−0.0854 to 0.0912).

**Figure 8.**
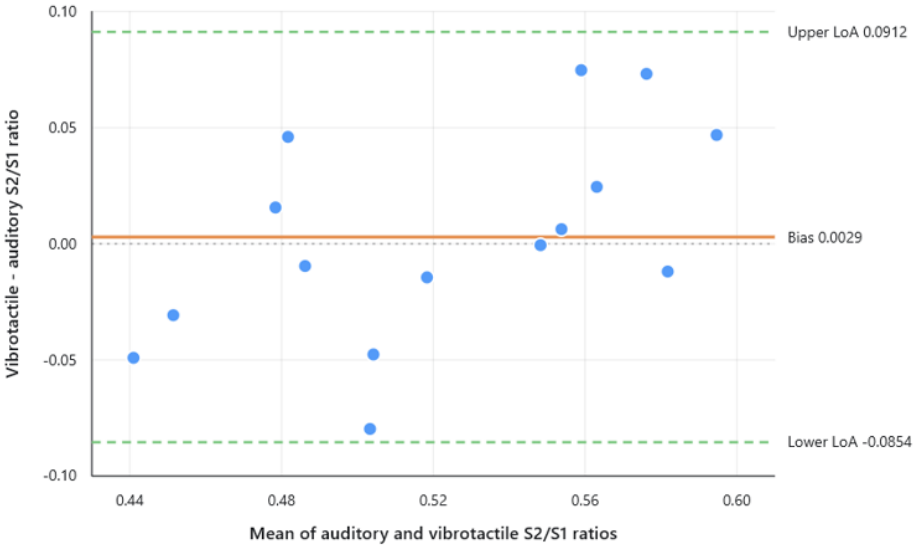
Bland-Altman plot comparing participant-level auditory and vibrotactile P50 S2/S1 ratios. Differences were calculated as vibrotactile minus auditory ratio. The solid orange line indicates the mean bias (0.0029), and the dashed green lines indicate the 95% limits of agreement (−0.0854 to 0.0912).

### B. Subject-Level P50 Ratios

Mean P50 S2/S1 ratio was 0.521 ± 0.041 for auditory stimulation and 0.524 ± 0.064 for vibrotactile stimulation. S2 was attenuated relative to S1 in both modalities.

The mean paired difference (vibrotactile minus auditory) was 0.0029 (95% CI, −0.0221 to 0.0279). No statistically significant difference was detected, t(14) = 0.249, p = 0.807, dz = 0.064.

Ratios were positively associated across participants (Pearson r = 0.715, p = 0.0027; Spearman ρ = 0.636, p = 0.0109), and ICC(A,1) was 0.665. Bland-Altman mean bias was 0.0029 with 95% limits of agreement from −0.0854 to 0.0912.

Paired differences had a median of −0.0006, IQR −0.0226 to 0.0353, and range −0.0797 to 0.0748. Regression suggested possible proportional bias (slope = 0.504, p = 0.035).

Median linear SNRs were 3.9 for auditory S1, 2.1 for auditory S2, 3.7 for vibrotactile S1, and 1.9 for vibrotactile S2.

| **Subject** | **Auditory Ratio** | **Vibrotactile Ratio** | **Vib - Aud Difference** |
|---|---|---|---|
| Subject 1 | 0.491 | 0.481 | -0.010 |
| Subject 2 | 0.551 | 0.557 | 0.006 |
| Subject 3 | 0.467 | 0.436 | -0.031 |
| Subject 4 | 0.540 | 0.613 | 0.073 |
| Subject 5 | 0.528 | 0.480 | -0.048 |
| Subject 6 | 0.549 | 0.548 | -0.001 |
| Subject 7 | 0.466 | 0.416 | -0.049 |
| Subject 8 | 0.588 | 0.576 | -0.012 |
| Subject 9 | 0.526 | 0.511 | -0.014 |
| Subject 10 | 0.543 | 0.463 | -0.080 |
| Subject 11 | 0.471 | 0.486 | 0.016 |
| Subject 12 | 0.551 | 0.575 | 0.025 |
| Subject 13 | 0.459 | 0.505 | 0.046 |
| Subject 14 | 0.571 | 0.618 | 0.047 |
| Subject 15 | 0.522 | 0.596 | 0.075 |

### C. Trial-Number Stability and Active Off-Skin Sham Control

Within the vibrotactile condition, no progressive trial-number trend was detected for S1 amplitude (slope = 0.00218 microvolts/trial, 95% CI -0.00219 to 0.00655, p = 0.303), S2 amplitude (slope = 0.00106 microvolts/trial, 95% CI -0.00134 to 0.00347, p = 0.359), or the S2/S1 ratio (slope = - 0.000018/trial, 95% CI -0.000378 to 0.000341, p = 0.914).

The trial-by-modality interactions were also nonsignificant for S1 (p = 0.216), S2 (p = 0.125), and the ratio (p = 0.426). Vibrotactile ratios were stable between trials 1-10 and trials 22-31 (0.5236 versus 0.5237; paired p = 0.973), with no early-to-late difference in S1 (p = 0.346) or S2 (p = 0.378). Figure S4 and Table S5 provide the complete sensitivity analysis.

In the active off-skin sham condition and electrical-coupling control, the powered transducer was operated adjacent to the forearm without skin contact. The P50 observed during skin-contact stimulation was not reproduced off the skin under the same acquisition and processing configuration.

### V. Discussion

Both conditions showed attenuation of S2 relative to S1. Mean auditory and vibrotactile ratios were 0.521 ± 0.041 and 0.524 ± 0.064, respectively, and the paired test detected no significant difference.

These findings are particularly noteworthy because the proposed system engages peripheral somatosensory pathways rather than the auditory system traditionally used in P50 paradigms. Although sensory gating has most frequently been investigated using auditory stimuli, the underlying inhibitory mechanisms are believed to reflect broader cortical processes involved in filtering repetitive sensory information. The observation that vibrotactile stimulation produced P50 attenuation of S2 responses supports the possibility that sensory-gating phenomena may extend across sensory modalities. While additional work is required to clarify the precise neural mechanisms involved, the present results provide preliminary evidence that peripheral somatosensory stimulation can serve as a viable substrate for sensory-gating assessment.

## VI. Limitations

This feasibility study included 15 healthy adults assessed during one recording session. The auditory condition preceded the vibrotactile condition in all participants; the approximately 10 s interval between successive paired-stimulus trials and the trial-number sensitivity analyses showed stable S1, S2, and S2/S1 measurements across all 31 trials. P50 amplitudes were quantified at Cz using a 128 samples/s portable system, and the Cz P50 waveforms and participant-level measurements are provided in the supplementary material. Figure S5 documents the EPOC Flex electrode montage. The separately supplied Fp1, Fp2, C3, and C4 workbook is labeled as auxiliary band-power data. The active off-skin sham condition and electrical-coupling control confirmed that the powered transducer did not reproduce the P50 without skin contact.

## VII. Conclusions

This laboratory feasibility study shows that portable EEG can record Cz P50 attenuation during vibrotactile paired stimulation in healthy adults. Further controlled validation is warranted.

Supplementary Materials: Table S1, Complete participant-level auditory and vibrotactile Cz P50 results; Figure S1, participant-level Cz waveforms; Table S2, participant-level auditory and vibrotactile Cz S1, S2, and S2/S1 values; Table S3, bootstrap confidence intervals; Table S4, participant-level SNR values; Figure S4, trial-number stability of Cz S1, S2, and S2/S1 measurements; Table S5, participant-adjusted trial-number and early-versus-late sensitivity analyses; Figure S5, EPOC Flex electrode montage; and the accompanying auxiliary band-power workbook for Fp1, Fp2, C3, and C4.

## Data Availability

All data produced in the present study are available upon reasonable request to the authors

## Author Contributions

Jean-Paul Badjo conceived and designed the study, developed the vibrotactile stimulation system and experimental methodology, collected the data, performed the EEG processing and formal analyses, interpreted the results, and prepared the original manuscript draft. Fow-Sen Choa contributed to the study design and interpretation of the results, supervised the research, provided guidance throughout the study, and critically reviewed and edited the manuscript. All authors have read and agreed to the published version of the manuscript.

## Institutional Review Board Statement

The study was conducted in accordance with the Declaration of Helsinki and approved by the Institutional Review Board of the University of Maryland, Baltimore County (protocol code 144 Y16FC37118 EEG-2 and date of approval Dec. 8, 2025).

## Informed Consent Statement

Informed consent was obtained from all subjects involved in the study. Written informed consent has been obtained from the participants to publish this paper.

## Data Availability Statement

The data presented in this study are available from the corresponding author upon reasonable request.

## Acknowledgments

The authors thank all study participants for their time and participation in this research.

## References

[1] Cromwell, H.C.; Mears, R.P.; Wan, L.; Boutros, N.N. Sensory gating: A translational effort from basic to clinical science. Clinical EEG and Neuroscience 2008, 39(2), 69–72. 10.1177/155005940803900209.

[2] Boutros, N.N.; Belger, A.; Campbell, D.; D’Souza, C.; Krystal, J. Comparison of four components of sensory gating in schizophrenia and normal subjects: A preliminary report. Psychiatry Research 1999, 88(2), 119–130. 10.1016/S0165-1781(99)00074-8.

[3] Patterson, J.V.; Hetrick, W.P.; Boutros, N.N.; Jin, Y.; Sandman, C.; Stern, H.; Potkin, S.; Bunney, W.E. P50 sensory gating ratios in schizophrenics and controls: A review and data analysis. Psychiatry Research 2008, 158(2), 226–247. 10.1016/j.psychres.2007.02.009.

[4] Light, G.A.; Braff, D.L. Sensory gating deficits in schizophrenia: Can we parse the effects of medication, nicotine use, and changes in clinical status? Clinical Neuroscience Research 2003, 3(1-2), 47–54. 10.1016/S1566-2772(03)00018-5.

[5] Potter, D.; Summerfelt, A.; Gold, J.; Buchanan, R.W. Review of clinical correlates of P50 sensory gating abnormalities in patients with schizophrenia. Schizophrenia Bulletin 2006, 32(4), 692–700. 10.1093/schbul/sbj050.

[6] Micoulaud-Franchi, J.-A.; Vaillant, F.; Lopez, R.; Peri, P.; Baillif, A.; Brandejsky, L.; Steffen, M.L.; Boyer, L.; Richieri, R.; Cermolacce, M.; et al. Sensory gating in adult with attention-deficit/hyperactivity disorder: Event-evoked potential and perceptual experience reports comparisons with schizophrenia. Biological Psychology 2015, 107, 16–23. 10.1016/j.biopsycho.2015.03.002.

[7] Sanchez-Morla, E.M.; Garcia-Jimenez, M.A.; Barabash, A.; Martinez-Vizcaino, V.; Mena, J.; Cabranes-Diaz, J.A.; Baca-Baldomero, E.; Santos, J.L. P50 sensory gating deficit is a common marker of vulnerability to bipolar disorder and schizophrenia. Acta Psychiatrica Scandinavica 2008, 117(4), 313–318. 10.1111/j.1600-0447.2007.01141.x.

[8] de la Salle, S.; Bowers, H.; Birmingham, M.; Phillips, J.L.; Blier, P.; Knott, V. Auditory P50 sensory gating alterations in major depressive disorder and their relationship to clinical symptoms. Psychiatry Research: Neuroimaging 2024, 341, 111813. 10.1016/j.pscychresns.2024.111813.

[9] Chien, Y.-L.; Hsieh, M.H.; Gau, S.S.-F. P50-N100-P200 sensory gating deficits in adolescents and young adults with autism spectrum disorders. Progress in Neuro-Psychopharmacology and Biological Psychiatry 2019, 95, 109683. 10.1016/j.pnpbp.2019.109683.

[10] Lijffijt, M.; Lane, S.D.; Meier, S.L.; Boutros, N.N.; Burroughs, S.; Steinberg, J.L.; Moeller, F.G.; Swann, A.C. P50, N100, and P200 sensory gating: Relationships with behavioral inhibition, attention, and working memory. Psychophysiology 2009, 46, 1059–1068. 10.1111/j.1469-8986.2009.00845.x.

[11] Du, X.; Hong, L.E. Test-retest reliability of short-interval intracortical inhibition and intracortical facilitation in patients with schizophrenia. Psychiatry Research 2018, 267, 575–581. 10.1016/j.psychres.2018.06.014.

[12] Wasaka, T.; Hoshiyama, M.; Nakata, H.; Nishihira, Y.; Kakigi, R. Gating of somatosensory evoked magnetic fields during the preparatory period of self-initiated finger movement. NeuroImage 2003, 20(3), 1830–1838. 10.1016/S1053-8119(03)00442-7.

[13] Hari, R.; Forss, N. Magnetoencephalography in the study of human somatosensory cortical processing. Philosophical Transactions of the Royal Society B 1999, 354, 1145–1154. 10.1098/rstb.1999.0470.

[14] Guidotti, M.; Beaurieux, C.; Marionnaud, P.; Bonnet-Brilhault, F.; Wardak, C.; Latinus, M. Skin type and nerve effects on cortical tactile processing: A somatosensory evoked potentials study. Journal of Neurophysiology 2023, 130(3), 547–556. 10.1152/jn.00444.2022.

[15] Jelincic, V.; D’Agostini, M.; Ventura-Bort, C.; Cascio, L.; Gorianskaia, E.; Weymar, M.; Torta, D.M.; Van Diest, I.; von Leupoldt, A. Continuous transcutaneous auricular vagus nerve stimulation increases long-latency neural processing in multiple sensory modalities. Psychophysiology 2025, 62(3), e70048. 10.1111/psyp.70048.

[16] Pesonen, H.; Strommer, J.; Li, X.; Parkkari, J.; Tarkka, I.M.; Astikainen, P. Magnetoencephalography reveals impaired sensory gating and change detection in older adults in the somatosensory system. Neuropsychologia 2023, 190, 108702.10.1016/j.neuropsychologia.2023.108702.

[17] Eidel, M.; Pfeiffer, M.; Ziebell, P.; Kubler, A. Recording the tactile P300 with the cEEGrid for potential use in a brain-computer interface. Frontiers in Human Neuroscience 2024, 18, 1371631.10.3389/fnhum.2024.1371631.

[18] Yang, J.; Ren, R.; Yu, Y.; Wang, W.; Tang, X.; Ejima, Y.; Wu, J. Event-related potential evidence for tactile orientation processing in the human brain. Experimental Brain Research 2024, 242(4), 809–817. 10.1007/s00221-024-06783-1.

[19] Schreiner, L.; Sieghartsleitner, S.; Cao, F.; Pretl, H.; Guger, C. Neural source reconstruction using a novel ultra-high-density EEG system and vibrotactile stimulation of individual fingers. In Proceedings of the 46th Annual International Conference of the IEEE Engineering in Medicine and Biology Society (EMBC), Orlando, FL, USA, 15-19 July 2024; pp. 1–5. 10.1109/EMBC53108.2024.10782167.

[20] Giannini, G.; Nierhaus, T.; Blankenburg, F. Investigation of sensory attenuation in the somatosensory domain using EEG in a novel virtual reality paradigm. Scientific Reports 2025, 15, 2819. 10.1038/s41598-025-87244-9.

[21] Debener, S.; Minow, F.; Emkes, R.; Gandras, K.; de Vos, M. How about taking a low-cost, small, and wireless EEG for a walk? Psychophysiology 2012, 49, 1617–1621. 10.1111/j.1469-8986.2012.01471.x.

[22] Badcock, N.A.; Mousikou, P.; Mahajan, Y.; de Lissa, P.; Thie, J.; McArthur, G. Validation of the Emotiv EPOC EEG gaming system for measuring research quality auditory ERPs. PeerJ 2013, 1, e38. 10.7717/peerj.38.

[23] Badcock, N.A.; Preece, K.A.; de Wit, B.; Glenn, K.; Fieder, N.; Thie, J.; McArthur, G.M. Validation of the Emotiv EPOC EEG system for research quality auditory event-related potentials in children. PeerJ 2015, 3, e907. 10.7717/peerj.907.

[24] Williams, N.S.; McArthur, G.M.; de Wit, B.; Ibrahim, G.; Badcock, N.A. A validation of Emotiv EPOC Flex saline for EEG and ERP research. PeerJ 2020, 8, e9713. 10.7717/peerj.9713.

[25] Epinat-Duclos, J.; Rossignon, A.; Prado, J.; Van der Henst, J.-B.; Paulignan, Y.; Beaudoin-Gobert, M.; Lecaignard, F.; Bedoin, N. Evaluating portable EEG: A comparison between two wireless systems (EPOC Flex and LiveAmp) and the wired BrainAmp system. PeerJ 2026, 14, e20416. 10.7717/peerj.20416.

[26] Williams, N.S.; McArthur, G.M.; Badcock, N.A. It’s all about time: Precision and accuracy of Emotiv event-marking for ERP research. PeerJ 2021, 9, e10700. 10.7717/peerj.10700.

[27] Mase, A.; Shibasaki, M.; Nakata, H. Effects of inter-stimulus and inter-trial intervals on somatosensory gating. Somatosensory & Motor Research 2025, 42(2), 55–60. 10.1080/08990220.2024.2358516.

[28] Delorme, A.; Makeig, S. EEGLAB: An open source toolbox for analysis of single-trial EEG dynamics including independent component analysis. Journal of Neuroscience Methods 2004, 134(1), 9–21.10.1016/j.jneumeth.2003.10.009.

[29] Luck, S.J. An Introduction to the Event-Related Potential Technique, 2nd ed.; MIT Press: Cambridge, MA, USA, 2014.

[30] Cohen, M.X. Analyzing Neural Time Series Data: Theory and Practice; MIT Press: Cambridge, MA, USA, 2014.

